# Human Milk Antibodies Elicited by BNT162b2 Vaccination have reduced activity against SARS-CoV-2 Variants of Concern

**DOI:** 10.1101/2021.09.20.21263808

**Authors:** Jia Ming Low, Yue Gu, Melissa Shu Feng Ng, Liang Wei Wang, Zubair Amin, Youjia Zhong, Paul A MacAry

## Abstract

We detected the presence of SARS-CoV-2 specific IgA against all major VOCs in milk out to 6 weeks after D2 of BNT162b2. These likely confer some protection to the breastfed infants, who are ineligible for vaccination and are at risk of severe COVID-19.

However, we detected significantly reduced milk IgA binding to VOCs, including the globally dominant Delta variant, suggesting reduced protection for breastfeeding infants. Additionally, these antibodies were significantly reduced by as early as 4-6 weeks after D2.

## Objective

SARS-CoV-2 specific antibody responses are engendered in human milk after BNT162b2 vaccination.^1^ However, the emergence of SARS-CoV-2 variants of concern (VOCs) raises questions about the specificity and potential cross-protection mediated by these antibodies. While antibody responses have been extensively studied for vaccinee sera,^2^ human milk antibodies – a major contributor to passive immunity for infants – have not been analyzed.

Here, we investigated whether BNT162b2 vaccination induced secretion of specific immunoglobulin A (IgA) into human milk against the principal determinant of neutralization (Spike-receptor binding domain, RBD) for 4 major VOCs. These data illuminate our understanding of transferred mucosal immunity in lactating mothers receiving mRNA vaccines.

## Study design

We conducted a prospective cohort study of lactating women. All received 2 doses of BNT162b2 (Pfizer/BioNTech) vaccine, given 21 days apart. Milk samples were collected pre-vaccination, 3-7 days after dose 2 (D2), and 4-6 weeks after D2. The study was approved by the Institutional Review Board (DSRB 2021/00095) and registered at ClinicalTrials.gov (NCT04802278); informed consent was obtained.

Enzyme-linked immunosorbent assay (ELISA) was used to assay IgA binding to SARS-CoV-2 RBD. IgA against Alpha (α, B.1.1.7), Beta (β, B.1.351), Gamma (γ, P.1), and Delta (δ, B.1.617.2) variants were tested using Kruskal-Wallis followed by Dunn’s Many-to-One Rank Comparison Test against the ancestral Wuhan-Hu-1 strain (WH-1). (See Supplemental Methods.)

## Results

46 subjects (mean age 31.5 years, 13.5 months post-partum) completed the study (Supplementary Table). IgA antibodies against RBDs of WH-1 strain, and 4 major VOCs were found in milk within 3-7 days after D2. Compared to WH-1, RBD-specific IgA was significantly reduced by 28-33% for the Beta, Gamma and Delta variants at this timepoint (Figure 1A). No difference was detected for the Alpha variant.

**Figure 1.**
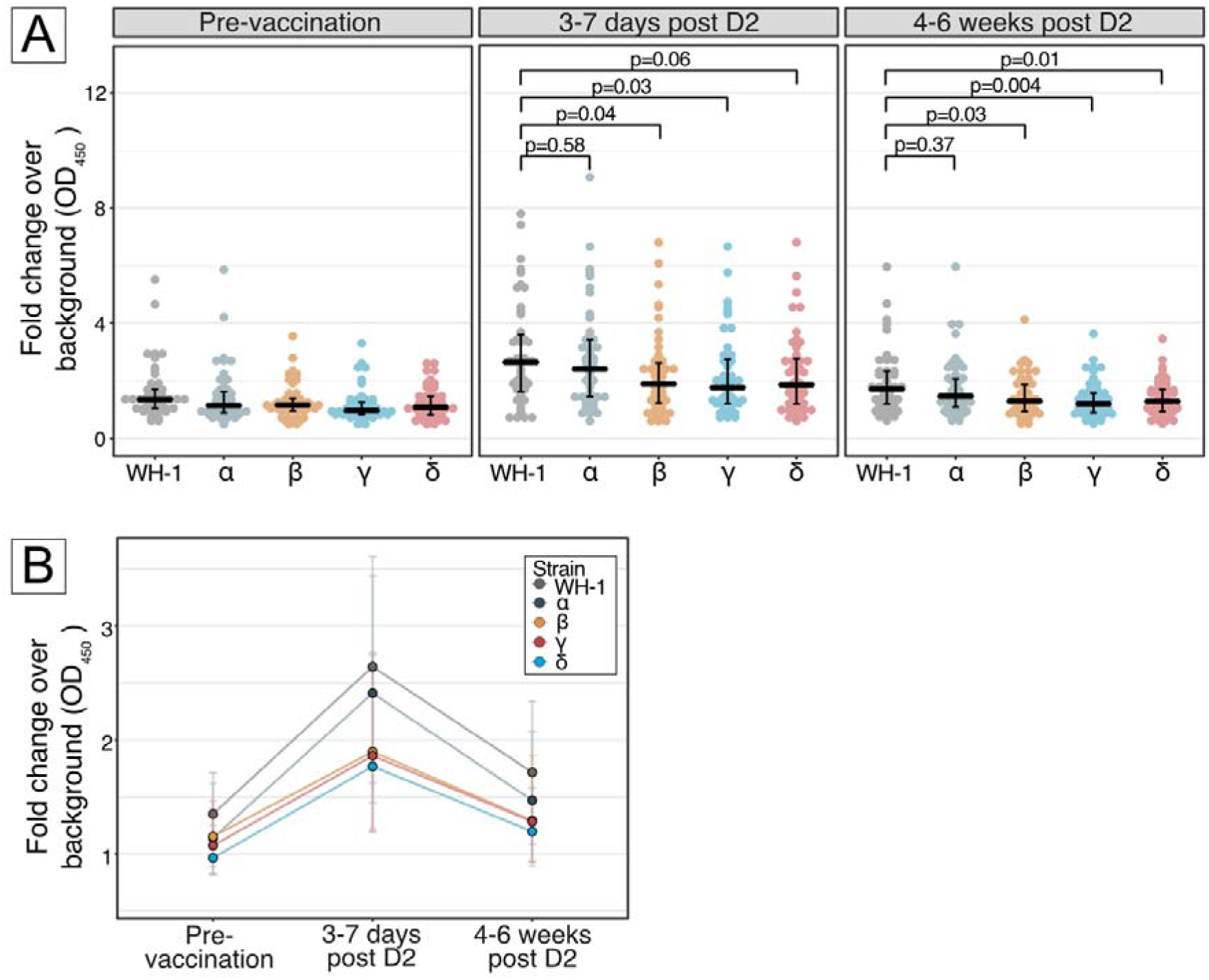
Detection of IgA binding to receptor binding domain of SARS-CoV-2 spike proteins encoded by the ancestral Wuhan-Hu-1 strain and selected major VOCs. **Panel A**, Differential human milk IgA binding to receptor binding domain (RBD) of SARS-CoV-2 spike protein of the ancestral Wuhan-Hu-1 strain (WH-1), Alpha (B.1.1.7), Beta (B.1.351), Gamma (P.1) and Delta (B.1.617.2) variants of concern, after vaccination with BNT162b2 across 3 time points: pre-vaccination, 3-7 days post D2 and 4-6 weeks post D2. Each dot denotes an individual sample. Centre line denotes the median, and error bars show Quartile 1 (bottom bar) and Quartile 3 (top bar). P values were calculated using Dunn’s Many-to-One Rank Comparison Test and reported for VOC compared against WH-1. At 3-7 days post D2, reduction from WH-1 for Alpha = 8%, Beta = 28%, Gamma = 33%, Delta = 30%. At 4-6 weeks post D2, reduction from WH-1 for Alpha = 14%, Beta = 25%, Gamma = 30%, Delta = 25%. **Panel B**, Kinetics of human milk IgA binding across 3 timepoints for ancestral Wuhan-Hu-1 strain (WH-1), Alpha (B.1.1.7), Beta (B.1.351), Gamma (P.1) and Delta (B.1.617.2) variants of concern. Each line represents 1 strain. Dots represent the median and error bars represent interquartile range.

At 4-6 weeks after D2, milk IgA against WH-1 and all VOCs decreased but were still higher than the pre-vaccination baseline. IgA binding relative to WH-1 was reduced for all VOCs except Alpha, by 25-30% (Figure 1B).

## Conclusion

We detected the presence of SARS-CoV-2 specific IgA against all major VOCs in milk out to 6 weeks after D2 of BNT162b2. These likely confer some protection to the breastfed infants, who are ineligible for vaccination and are at risk of severe COVID-19. ^3^

However, we detected significantly reduced milk IgA binding to VOCs, including the globally dominant Delta variant, suggesting reduced protection for breastfeeding infants. Additionally, these antibodies were significantly reduced by as early as 4-6 weeks after D2^1^.

This study lends supports to the increasing importance of antenatal vaccination of pregnant women to facilitate transplacental transfer of IgG to the infant as it wanes slower at 5-6 months of life thereby augmenting both the magnitude and duration of passive immunity transferred from mother to child.^4^ Additionally, lactating mothers as a population could be prioritized for booster vaccine doses to maximize transferred immunity to vulnerable infants.

We acknowledge certain limitations of this study. No functional assays were performed; however, SARS-CoV-2 specific IgA binding in vaccinee milk has been positively correlated with neutralization.^5^ This is a small and relatively homogenous population; larger studies are warranted.

## Data Availability

The data can be made available upon request.

## Acknowledgements

J.M.L. received funding from KTP—NUCMI and NUS Yong Loo Lin School of Medicine Pitch For Funds Grant to conduct this research. M.S.F.N. is a recipient of the Career Development Award from the Agency for Science, Technology and Research, Singapore. L.W.W. is a recipient of the National Medical Research Council Open Fund-Young Investigator Research Grant (Project ID: MOH-000545-00) from the Ministry of Health, Singapore. This study is funded by the SARS-CoV-2 antibody initiative (R-571-000-081-213) and Reimagine research fund (R-571-001-093-114). The authors wish to thank Miss Regena Chua Xin Yi (BsC) for helping with data collection and Dr Dimple Rajgor (PhD) for helping with preparing the manuscript for publication. We thank Prof Paul Anantharajah Tambyah for provision of funding and Dr Le Ye Lee (MRCPCH) for providing mentorship. No compensation was received for their roles. Last but not least, we are deeply grateful to all mothers who donated their time and precious gift of milk to science.

## Supplemental methods

We conducted a prospective cohort study of a convenience sample of lactating women in Singapore. Participants were recruited between February 5-9, 2021 through advertisements. All received 2 doses of BNT162b2 (Pfizer/BioNTech) vaccine, with the second dose (D2) given 21 days after dose 1. Human milk samples were collected before administration of vaccine, 3-7 days after D2, and 4-6 weeks after D2. The study was approved by the Institutional Review Board (DSRB 2021/00095); informed consent was obtained. The study was registered at ClinicalTrials.gov (NCT04802278).

Milk samples were delipidated by centrifugation twice, at 10000g, 4 °C for 15 minutes each time. The clear portion was stored at -20 °C and thawed before use.

Receptor binding domain (RBD) of SARS-CoV-2 ancestral Wuhan-Hu-1 strain (WH-1), Alpha (B.1.1.7), Beta (B.1.351), Gamma (P.1) and Delta (B.1.617.2) variants were diluted in PBS and coated on 384-well Maxisorp plates (NUNC) at 80 ng/well for overnight incubation at 4 °C. Plate was washed 3 times with 1x PBST buffer (0.05% Tween in 1x PBS) and blocked with 3% bovine serum albumin in 1x PBST for 1.5 hour incubation at room temperature. Delipidated milk was diluted 20-fold in the blocking buffer. After 3 plate washes in 1x PBST, diluted milk samples were added at 20 µL/well for 1 hour incubation at room temperature. Plate wash procedure was repeated thrice. Anti-human F(ab’)2 anti-human IgA-HRP (Invitrogen, #A24458) was diluted 5000-times in the blocking buffer and added to the plate at 20 µL/well for 1 hour incubation at room temperature, protected from light. After 3 plate washes in 1x PBST, 1-Step Ultra TMB-ELISA (Thermo Scientific, #34029) was added at 20 µL/well. Reaction was stopped with 20 µL/well 1 M H_2_SO_4_ 3 minutes later. OD_450_ was measured using a microplate reader (Tecan Spark). Results were based on 3 technical replicates per sample.

Statistical analysis was performed with R (4.0.2) using the PMCMRPlus package (1.9.0). Tests performed were Kruskal-Wallis followed by Dunn’s Many-to-One Rank Comparison Test for comparisons of variants to WH-1

**Supplementary Table.** Characteristic of lactating women.

| **Supplementary Table. Characteristic of lactating women** |  |
| --- | --- |
|  | **No. (%)** |
| Study participants, No. | 46 |
| **Maternal characteristics** |  |
| Maternal Age, mean [standard deviation, SD], years | 31.5 [0.9] |
| Ethnicity |  |
| Singaporean Chinese | 41 (89.1) |
| Singaporean Malay | 5 (10.9) |
| Chronic diseases |  |
| Asthma | 1 (2.2) |
| Polycystic Ovarian Syndrome | 1 (2.2) |
| Thalassemia Minor | 1 (2.2) |
| Antenatal conditions | 0 |
| Smoker | 0 |
| Postpartum [standard deviation, SD], months | 13.5 [SD 2.1] |

